# Assessing drug-mediated inhibition of liver transporter function with MRI: A first-in-human study

**DOI:** 10.1101/2025.06.16.25329670

**Authors:** Thazin Min, Marta Tibiletti, Paul D Hockings, Aleksandra Galetin, Ebony Gunwhy, Eve Shalom, J. Gerry Kenna, Nicola Melillo, Geoff JM Parker, Gunnar Schuetz, Daniel Scotcher, John C Waterton, Ian Rowe, Steven Sourbron

**Author notes:** Thazin Min and Marta Tibiletti contributed equally to this work. Ian Rowe and Steven Sourbron are co-senior authors.

## Abstract

**Background:** Assessing the risk of drug-mediated liver injury (DILI) and liver-mediated drug-drug interactions (DDI) for new drugs requires biomarkers that respond to drug effects on hepatic transporters. Evidence in rats has shown that dynamic gadoxetate-enhanced MRI (DGE-MRI) is suitable for this purpose, but it is not known whether these findings translate to humans.

**Purpose:** To determine the effects of rifampicin, a known inhibitor of hepatocyte transporters OATP1B and MRP2, on gadoxetate uptake and excretion rates in the liver of healthy volunteers.

**Materials and Methods:** This prospective study recruited 10 healthy volunteers, who were assessed on two separate visits. DGE-MRI was performed over two separate scans, one hour apart, due to the slow hepatic excretion of gadoxetate after inhibition. DGE was acquired with a fast (2.5 sec) 3D free-breathing protocol collecting data continuously for 50 minutes. Gadoxetate was injected at 1/8^th^ of a clinical dose, escalated to 1/4^th^ after the first 3 volunteers. On the second visit, rifampicin (600mg) was administered orally one hour before the start of the scan. Liver uptake and excretion rates were derived by modelling signal-time curves in aorta and liver. The effect of rifampicin was determined with a paired t-test with significance level at p<0.01.

**Results:** Eight of the 10 participants (3 female/7 male, mean age 32) completed both visits. Rifampicin reduced hepatocellular uptake rate of gadoxetate by 93% (95%CI 91-95%, p<0.001). Biliary excretion rate was reduced by 50% (p=0.004) but the effect was more variable (95%CI 8- 92%). Both rates were reduced by rifampicin in every participant, except for the excretion rate of one participant who had low baseline levels.

**Conclusion:** MRI measurements of gadoxetate biliary excretion and liver uptake rates can robustly detect inhibition of hepatocellular function mediated via OATP1B and MRP2 transporters.

## Introduction

Drug induced liver injury (DILI) can be caused by numerous drugs in humans and may result in a spectrum of clinical presentations from asymptomatic to life-threatening acute liver failure [1,2]. DILI may present as hepatocellular (i.e. affecting hepatocytes), cholestatic (affecting bile flow and the biliary system) and mixed. Hepatobiliary transport inhibition by drugs is thought to be one of the main mechanisms related to drug-induced cholestasis [3]. Numerous clinically relevant drug-drug interactions (DDIs) also arise via inhibition of hepatic uptake or excretion of drugs, which may affect efficacy and toxicity [4]. Drug-mediated transporter inhibition is currently assessed by a combination of in vitro and in silico methods [5]. Unfortunately, model-based predictions of changes in liver exposure are difficult to verify clinically, especially when the drug inhibits biliary excretion [6].

Reducing the risk of DILI and DDIs by novel drugs therefore requires a more accurate measurement of inhibition of liver uptake and excretion by drugs in humans.

A potential solution is provided by dynamic gadoxetate-enhanced MRI (DGE-MRI). Gadoxetate is a substrate of the liver uptake and biliary efflux transporters OATP1B1, OATP1B3, NTCP, MRP2 and MRP3 [7,8,9], and is approved as a radiographic contrast agent to detect and characterize liver lesions [10]. With appropriate modelling and analysis, the data can be used to quantify gadoxetate liver uptake- and excretion rates in humans [11,12]. In a context of DDI prediction, these data can then be combined with preclinical- and in vitro data to predict the effect of transporter inhibitor drugs on the hepatic disposition more accurately [13].

Studies in rats have shown that inhibition of liver uptake and excretion caused by drugs can be measured reproducibly with DGE-MRI, can distinguish inhibitors from non-inhibitors, and inhibition of hepatocellular uptake from inhibition of biliary excretion [9,14,15]. Based on this evidence the Food and Drug Administration (FDA) has accepted the gadoxetate-MRI assay into their biomarker qualification program [16], but data on efficacy in humans is critically needed to evidence utility in drug development and basic research.

The aim of this study was to test whether findings in rats translate to humans, by measuring the change in liver gadoxetate uptake and excretion after administration of a known potent and selective inhibitor drug in healthy volunteers. Rifampicin was selected as a test drug since the preclinical studies revealed a strong inhibition of biliary efflux in addition to the expected inhibition of OATP1B activity.

## Materials and Methods

### Study design

The study was approved by the local research ethics committee and prospectively recruited 10 healthy volunteers, who gave informed consent. Exclusion criteria were previous history of liver or kidney disease and regular prescribed medication, including oral contraceptives.

After informed consent, each participant underwent two study visits on different days, 2-4 weeks apart (Fig 1). The visits were identical except for an extra blood sample and oral administration of 600 mg rifampicin on visit 2, one hour before the start of MRI. Timing of administration was based on simulations aiming for maximum inhibition at the start of DGE-MRI [9,13].

**Figure 1:**
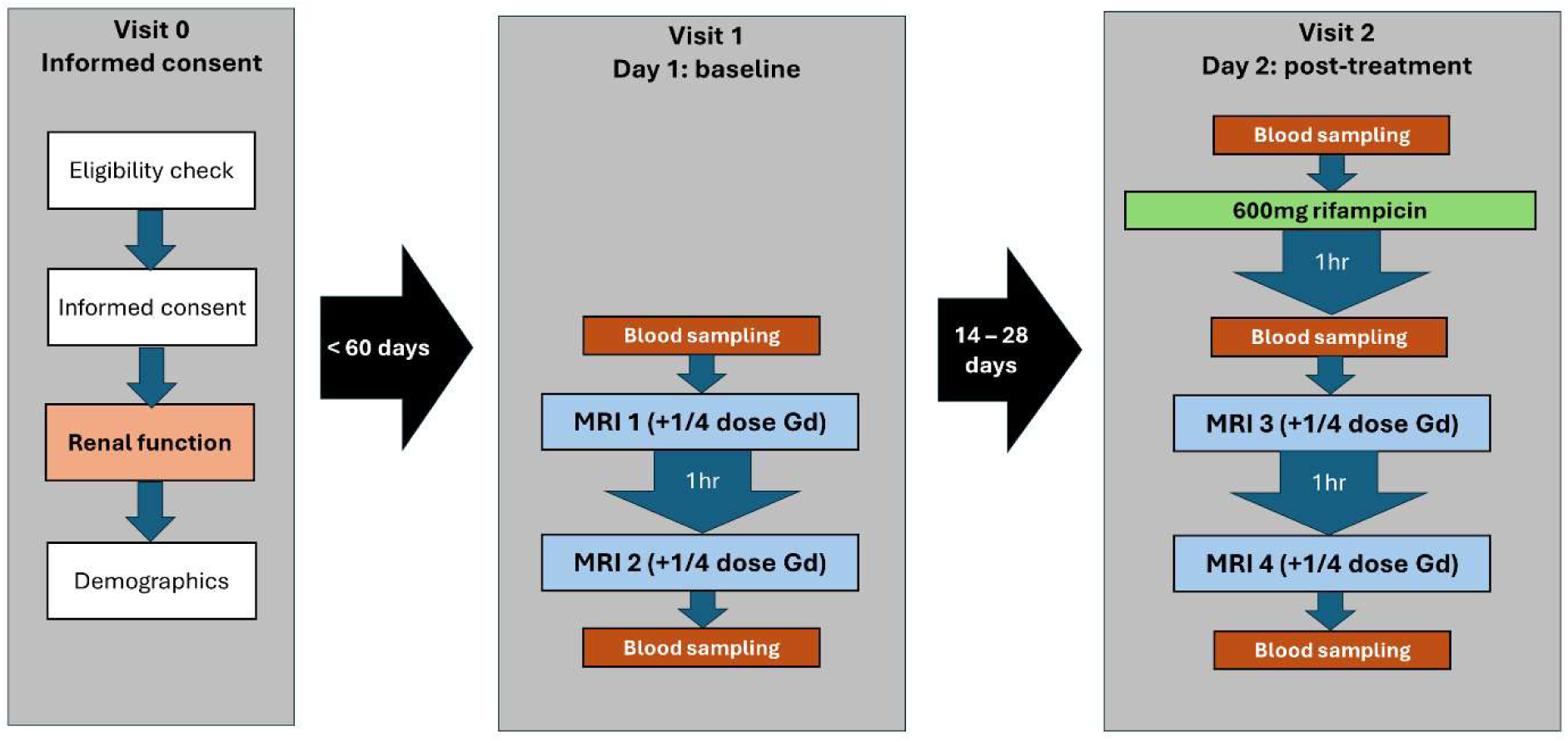
Flow diagram of the final protocol after dose escalation, showing the study design and assessments performed at each visit. Visit 0 was a screening visit to assess eligibility for the study, take informed consent and collect key data. Visit 1 was the baseline visit, no more than 60 days after consent. It involved two MRI scans at least 2 hours apart, and blood tests before and after the scans. The post-treatment visit 2 took place between 2 and 4 weeks after the baseline visit, and involved blood sampling before oral rifampicin administration, then after 1 hour a repeat of the same protocol performed at visit 1.

Since biliary excretion of gadoxetate after inhibition is slow, the measurement on each day was split over two scans, separated by an hour where the participant left the scanner and could rest. In each of the two scans, DGE-MRI was performed for 50 minutes. Participants were allowed to eat and drink before the study in moderation. Blood samples were taken before and after each scan for standard liver function tests (LFT’s).

The study design included a gadoxetate dose escalation strategy to ensure that no higher gadoxetate dose was used than strictly necessary. Initial dosage was one eighth of a standard clinical dose in each of the four scans. The data were reviewed after two participants completed both visits and the protocol allowed an increment in the dose at this stage.

### MRI protocol

MRI was performed in a single site on a 3T Siemens Prisma. The protocol included localizers, high-resolution T2-weighted axial and coronal scans, T2* mapping, transverse T1-mapping with a modified Look-Locker sequence (MOLLI), B1-mapping, coronal variable flip angle T1- mapping (VFA) and coronal free-breathing DGE-MRI. The first MRI of each day concluded with a post-contrast transverse MOLLI T1 measurement. T2*, B1 and VFA T1 were acquired for secondary objectives and are not reported in this paper.

MOLLI was acquired with 5 slices, 5 mm thick, FA 12 deg, TR/TE 423/2.75 ms, 7(2)4 scheme with simulated ECG trace at 80 bpm. The DGE sequence was a 3D FLASH with a time resolution of 2.3 s, 36 slices, FOV 450 mm, TR 4.93 ms, TE 1.23 ms, flip angle 15 deg, voxel size 4.7x4.7x5 mm^3^, base resolution 96x96 and 3D GRAPPA factor 2. Gadoxetate was injected at 1 mL/s and followed by a 20 mL saline flush at the same rate. Injection was performed 5 min after the start of the DGE-MRI sequence on the first scan, and after 10 min on the second.

### MRI preprocessing

Anonymized DICOM images were uploaded on a central server and processed using 21CFR11-compliant software, Voxelflow (Bioxydyn, Manchester UK). The MOLLI T1 maps calculated by the scanner software (syngo MR E11) were manually segmented to extract median whole liver and aorta T1 values. Whole liver regions were drawn avoiding major blood vessels. Breathing motion and patient positioning between scans was corrected using 3D rigid and deformable motion correction [17]. Signal-time curves for both scans were exported as csv files for DGE-MRI modelling [18].

### DGE-MRI signal processing

Signal processing was performed by freely available python scripts [19]. MRI modelling was performed with the open-source package dcmri.org, using the function AortaLiver2scan [20]. Gadoxetate concentrations in the aorta are modelled using a whole-body model of the circulation, then used as input to a two-site liver model with uptake into hepatocytes, and excretion into bile. Uptake and excretion rates k(he) and k(bh) were allowed to vary linearly over time to allow for changes in liver function during the day. Concentrations were converted to longitudinal relaxation rates using the precontrast MOLLI T1 and literature values for the gadoxetate relaxivities in liver and in blood at 3T [22]. Relaxation rates were converted to signals using the standard model for a spoiled gradient echo sequence in the steady state [23]. The model was fitted to the signals using default initial values and constraints for all parameters.

The analysis produced the primary hepatocellular (HC) outcome markers, biomarkers characterizing the extracellular space (EC), and simpler semi-quantitative (SQ) measures for comparison - such as R1 (=1/T1) and signal-enhancement at 20 minutes. For comparison with non-imaging pharmacokinetic studies, MRI-measured liver volumes were used to derive liver blood clearance (CL, mL/min).

### Data analysis

Statistical analysis including generation of tables and figures was performed by freely available python scripts [21].

Effect sizes were calculated for each participant and each parameter as the relative difference from baseline value. Mean values and 95% confidence intervals on the mean (95%CI) were calculated for all parameters at baseline and after rifampicin, and for their effect sizes. A two-sided paired t-test was used to evaluate differences between baseline and rifampicin values. Effect sizes of parameters were compared using the t-statistic (t) and a correlation analysis was performed on baseline values and absolute effect sizes to identify clusters of similar biomarkers. p<0.01 was considered significant for all tests.

The primary outcome markers were the hepatocellular uptake rate k(he) and biliary excretion rate k(bh) at the mid-point of the acquisition. Secondary DGE-MRI markers were also tested for rifampicin effect as exploratory outputs. For comparison between MRI and LFT’s, the pre-scan and post-scan LFT values were averaged.

## Results

Ten volunteers (3 female, 7 male) were recruited and underwent the baseline scan (Table 1). LFT’s before the first scan verified that all participants had normal liver function, and that liver function did not change between visits (Table 2). No incidental imaging findings were noted for any of the 10 participants.

**Table 1:**
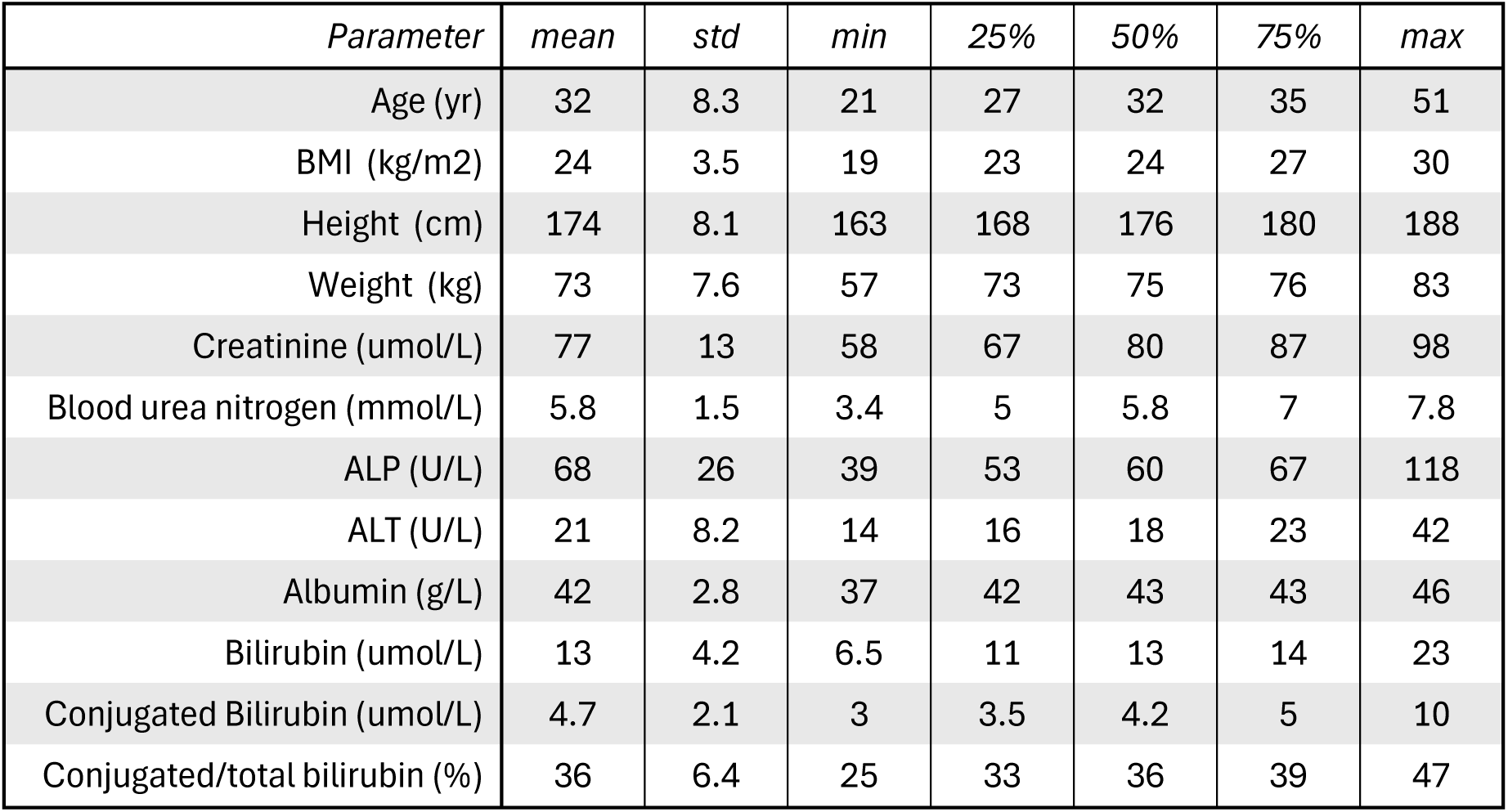
Demographics and clinical characteristics of the 10 participants recruited into the study. 3 of the participants (30%) were female.

| *Parameter* | *mean* | *std* | *min* | *25%* | *50%* | *75%* | *max* |
| --- | --- | --- | --- | --- | --- | --- | --- |
| Age (yr) | 32 | 8.3 | 21 | 27 | 32 | 35 | 51 |
| BMI (kg/m2) | 24 | 3.5 | 19 | 23 | 24 | 27 | 30 |
| Height (cm) | 174 | 8.1 | 163 | 168 | 176 | 180 | 188 |
| Weight (kg) | 73 | 7.6 | 57 | 73 | 75 | 76 | 83 |
| Creatinine (umol/L) | 77 | 13 | 58 | 67 | 80 | 87 | 98 |
| Blood urea nitrogen (mmol/L) | 5.8 | 1.5 | 3.4 | 5 | 5.8 | 7 | 7.8 |
| ALP (U/L) | 68 | 26 | 39 | 53 | 60 | 67 | 118 |
| ALT (U/L) | 21 | 8.2 | 14 | 16 | 18 | 23 | 42 |
| Albumin (g/L) | 42 | 2.8 | 37 | 42 | 43 | 43 | 46 |
| Bilirubin (umol/L) | 13 | 4.2 | 6.5 | 11 | 13 | 14 | 23 |
| Conjugated Bilirubin (umol/L) | 4.7 | 2.1 | 3 | 3.5 | 4.2 | 5 | 10 |
| Conjugated/total bilirubin (%) | 36 | 6.4 | 25 | 33 | 36 | 39 | 47 |

**Table 2:** Liver function tests (LFT) at the start of each visit showing no systematic difference in liver function between visits. For each parameter the table shows mean values (95% on the mean) at the start of visit 1 (Baseline) and visit 2 (Rifampicin), the difference between both and the p-value for the paired t-test.

| *Parameter* | *Visit 1* | *Visit 2* | *Diff* | *p-value* |
| --- | --- | --- | --- | --- |
| ALP (U/L) | 68 (40) | 69 (54) | 1.8 (23) | 0.69 |
| ALT (U/L) | 22 (19) | 25 (19) | 3.6 (9.4) | 0.07 |
| Albumin (g/L) | 44 (4.1) | 44 (5.9) | -0.6 (6.3) | 0.6 |
| Bilirubin (umol/L) | 12 (8.5) | 11 (9.3) | -0.8 (12) | 0.73 |
| Conjugated Bilirubin (umol/L) | 4.8 (3.7) | 4.6 (4.2) | -0.1 (5.1) | 0.89 |
| Conjugated/total bilirubin (%) | 39 (11) | 37 (9.1) | -3.0 (16) | 0.36 |

Eight volunteers completed all visits. In one volunteer, the gadoxetate at baseline was mistakenly administered at 1.25 times a standard clinical dose instead of 0.125 as prescribed. The volunteer was withdrawn from the study and followed up clinically, but no adverse events were reported. Another participant could not complete the second visit because of difficulties with venous access.

Following the staged design, data from the first two complete baseline/rifampicin studies were reviewed before continuing. At this point it was decided to escalate the gadoxetate dose per scan from 1/8th to 1/4th of a clinical dose to boost the enhancement-to-noise level. Data for both doses were combined for analysis.

Visual inspection of signals (Fig 2) show that the model fits the data well and reveals clear differences after rifampicin, consistent with reduced uptake and delayed excretion. At baseline the extracellular phase of the signal is masked by a rapid uptake which peaks after approximately 25 minutes (Fig 2a). After rifampicin, uptake is much reduced with no obvious hepatobiliary peak concentration, revealing the extracellular phase (Fig 2b). The second injection shows a similar pattern as the first. Visually, the aorta signal at baseline and after rifampicin is similar (Fig 2 c, d).

**Figure 2:**
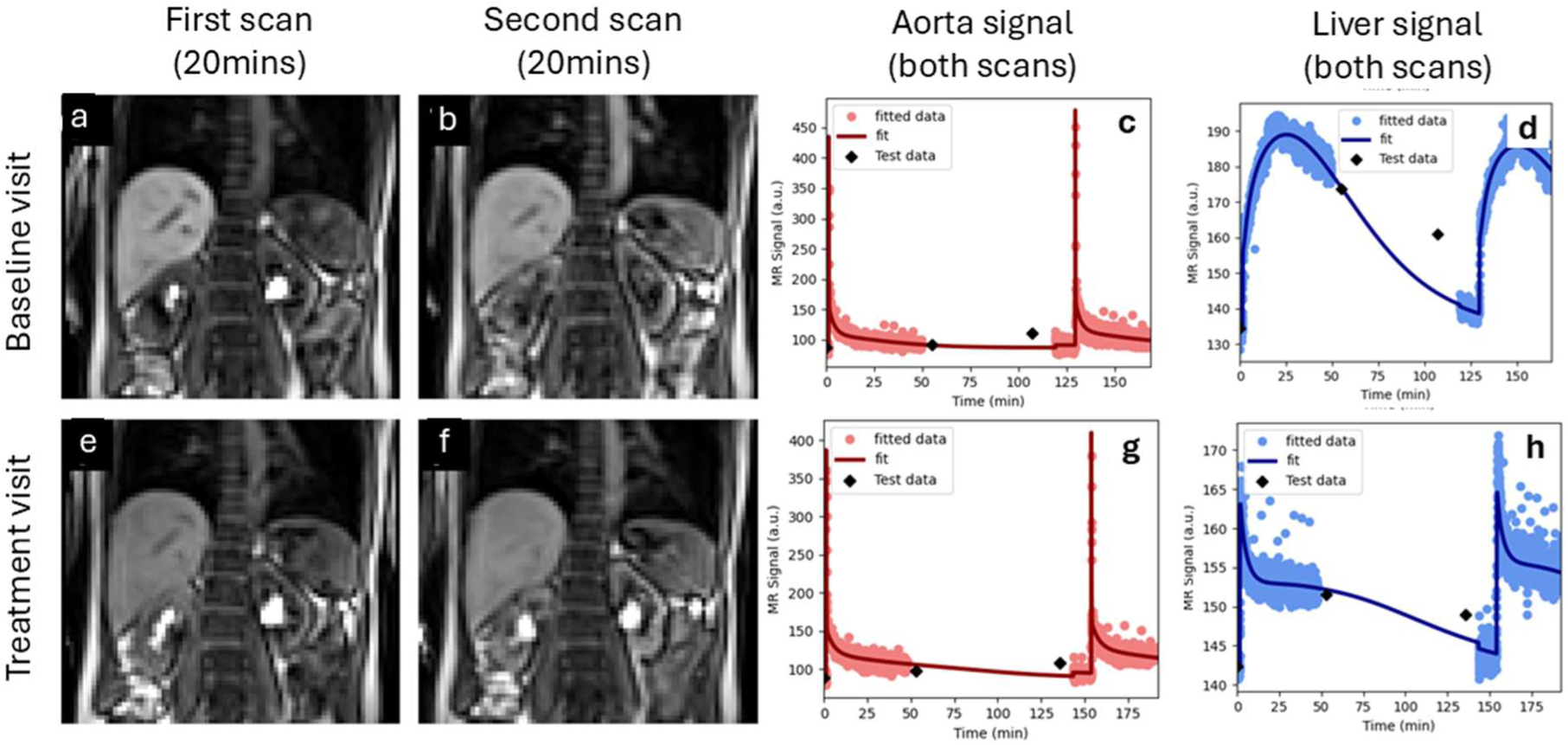
Signals for the two scans performed on the same day for one volunteer at baseline (top row) and after a single 600 mg dose of rifampicin (bottom row). The top row shows an image of the first scan (a) and second scan (b) at peak enhancement (20mins after injection), and signal-time curves of gadoxetate data in the aorta (c) and liver (d). The bottom row (e-h) shows the same data for the treatment visit. The plots shows the signal with the model fit superposed, showing a small discontinuity before the second scan due to signal recalibration. The MOLLI T1-values are fed through the signal model and plotted as part of the quality control process (checkpoints) but are not used in the actual model fit.

Table 3 shows numerical results for all liver biomarkers. On average, rifampicin reduces the hepatocellular uptake rate k(he) by 93% (95%CI 91-95%, p<0.001). The biliary excretion rate k(bh) reduces by only 50% and the effect is more variable (95%CI 8-92%, p=0.004). The liver blood clearance shows the strongest response (t = 15); most semi-quantitative (Type=SQ) measures also show a clear effect (4.5 < t < 12).

**Table 3.** Drug effect on the liver biomarkers including, in the top two rows, the primary outcome markers k(he) (hepatocellular uptake rate) and k(bh) (biliary excretion rate). The first column is the type of biomarker (HC = Hepatocellular, SQ = semi-quantitative, EC = extracellular). The other columns show mean (95% CI on the mean) values at baseline and after rifampicin administration, their relative effect size as well as the t-statistic and p-value for a paired t-test. Biomarkers that showed a significant change after rifampicin (p<0.01) are highlighted in bold.

| *Short name - parameter (units)* | *Type* | *Baseline* | *Rifampicin* | *Effect (%)* | *t* | *p* |
| --- | --- | --- | --- | --- | --- | --- |
| **k(he) - Hepatocellular uptake rate (mL/min/100cm<sup>3</sup>)** | **HC** | **30 (4.5)** | **2.2 (0.6)** | **-93 (2.5)** | **11** | **<0.001** |
| **k(bh) - Biliary excretion rate (mL/min/100cm<sup>3</sup>)** | **HC** | **2.2 (0.5)** | **0.79 (0.34)** | **-49 (42)** | **4.1** | **0.005** |
| **Cl - Liver blood clearance (mL/min)** | **HC** | **265 (28)** | **20 (4)** | **-92 (2.5)** | **15** | **<0.001** |
| **RES(l) - Relative enhancement at 20 min (%)** | **SQ** | **66 (23)** | **6.5 (1.7)** | **-87 (1.9)** | **8.8** | **<0.001** |
| **RER1(l) - Relative R1 enhancement at 20 min (%)** | **SQ** | **87 (40)** | **8.9 (1.5)** | **-86 (1.9)** | **8.3** | **<0.001** |
| **AUC(30,l) - Area under the curve at 30 min (mM\*sec)** | **SQ** | **205 (100)** | **26 (4.3)** | **-82 (2.8)** | **8.6** | **<0.001** |
| **R1(45) - R1 at 45 min (1/sec)** | **SQ** | **2.3 (0.8)** | **1.4 (0.08)** | **-26 (3.2)** | **12** | **<0.001** |
| **DR1(45) - R1change at 45 min (1/sec)** | **SQ** | **1.1 (0.8)** | **0.14 (0.03)** | **-77 (5.8)** | **8.7** | **<0.001** |
| **R1(2) - R1 at scan 2 (1/sec)** | **SQ** | **1.7 (0.4)** | **1.3 (0.07)** | **-12 (4.6)** | **4.5** | **0.003** |
| DR1(2) - Change in R1 at scan 2 (1/sec) | SQ | 0.46<br>(0.43) | 0.08 (0.02) | -31 (76) | 3.1 | 0.02 |
| v(e) - Extracellular volume (mL/100cm<sup>3</sup>) | EC | 18 (6.5) | 22 (3.4) | 264 (420) | -1.4 | 0.2 |
| MTT(e) - Extracellular mean transit time (sec) | EC | 36 (11) | 43 (7.7) | 148 (240) | -0.9 | 0.41 |
| TTD(e) - Extracellular transit time dispersion (%) | EC | 66 (16) | 71 (4.8) | 283 (550) | -0.7 | 0.52 |

Table 4 shows results for all LFT’s. Bilirubin shows a significant change of 82% (95%CI 30- 134, p=0.001), but the effect is substantially weaker than most MRI biomarkers (t = -5.4). The other LFT’s show no systematic effect.

**Table 4:**
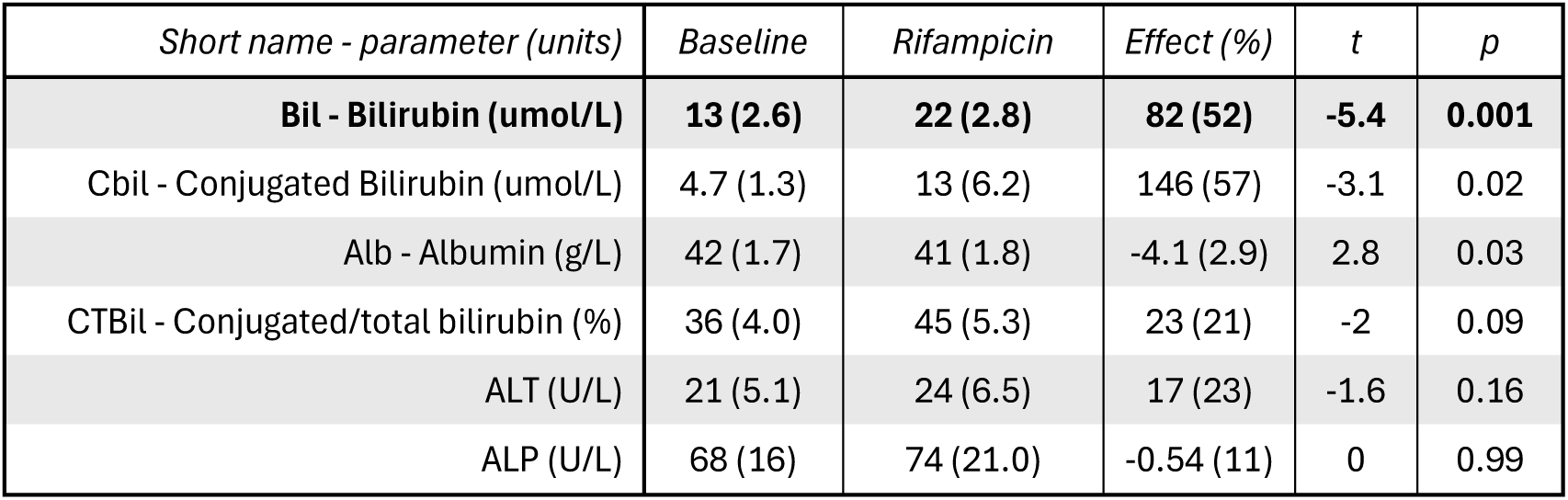
Drug effect on LFT’s, showing mean (95% CI on the mean) values at baseline and after rifampicin administration, their relative effect size (%) as well as the t-statistic and p- value for a paired t-test. Biomarkers that showed a significant change after rifampicin (p<0.01) are highlighted in bold.

| *Short name - parameter (units)* | *Baseline* | *Rifampicin* | *Effect (%)* | *t* | *p* |
| --- | --- | --- | --- | --- | --- |
| **Bil - Bilirubin (umol/L)** | **13 (2.6)** | **22 (2.8)** | **82 (52)** | **-5.4** | **0.001** |
| Cbil - Conjugated Bilirubin (umol/L) | 4.7 (1.3) | 13 (6.2) | 146 (57) | -3.1 | 0.02 |
| Alb - Albumin (g/L) | 42 (1.7) | 41 (1.8) | -4.1 (2.9) | 2.8 | 0.03 |
| CTBil - Conjugated/total bilirubin (%) | 36 (4.0) | 45 (5.3) | 23 (21) | -2 | 0.09 |
| ALT (U/L) | 21 (5.1) | 24 (6.5) | 17 (23) | -1.6 | 0.16 |
| ALP (U/L) | 68 (16) | 74 (21.0) | -0.54 (11) | 0 | 0.99 |

Figure 3 shows the individual results for the primary outcomes. Both rates reduce in each participant with the exception of one k(bh) value which is low at baseline (Fig3 c, red line). Visual assessment confirmed atypical kinetics in that participant showing no peak enhancement within the acquisition window (not shown). The participant was the oldest in the group by 13 years and had the highest BMI (30).

**Figure 3.**
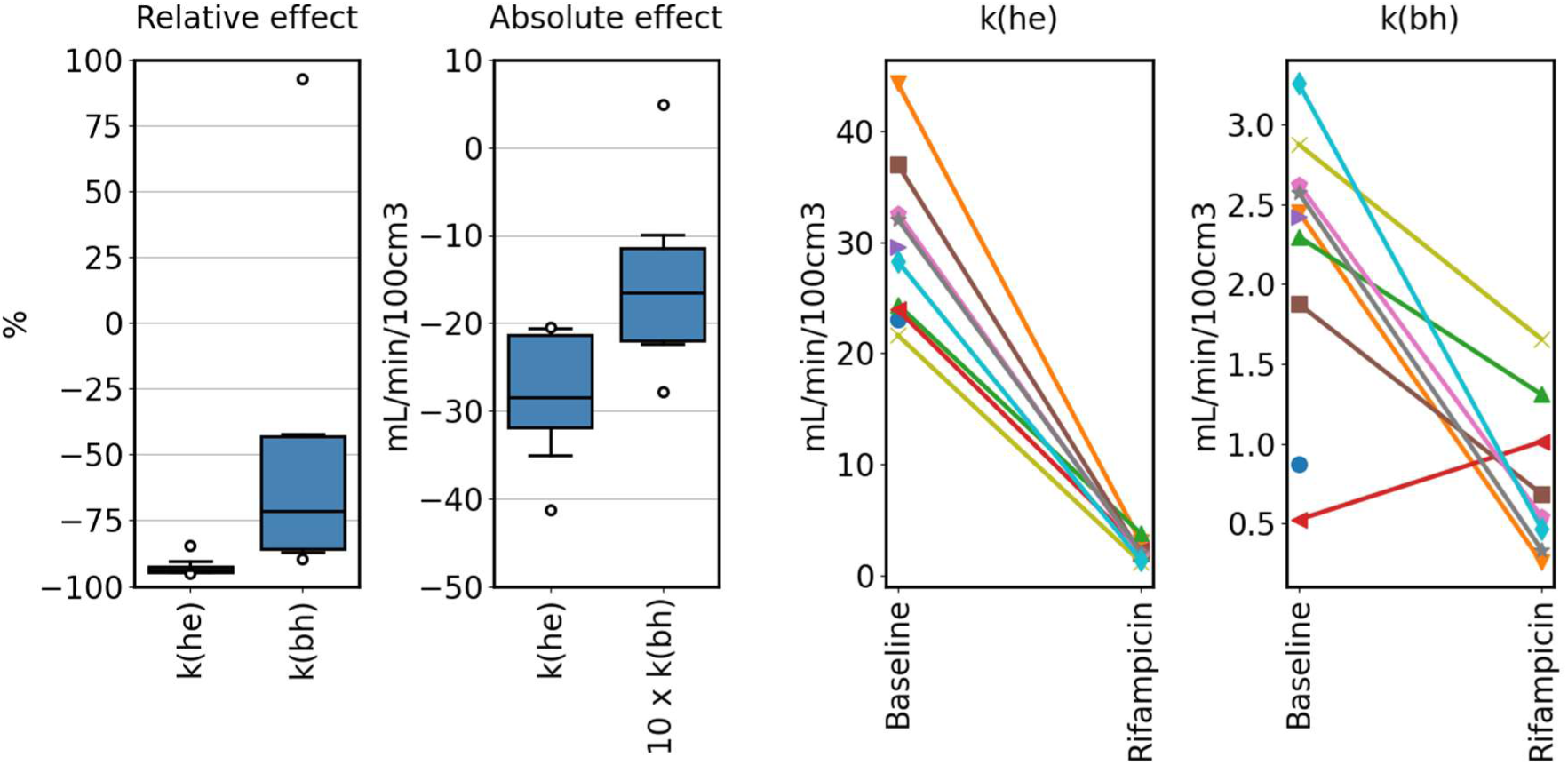
Rifampicin effect on the primary endpoints k(he) (hepatocellular uptake rate) and k(bh) (hepatocellular excretion rate) across the population. Box plots on the left show the relative and absolute effect size across the population. Absolute effect sizes for k(bh) have been scaled with a factor 10 to improve visualisation. Line plots on the right show individual values at baseline and after single dose of rifampicin, with each line representing an individual participant. Values for participants that attended the baseline visit only are shown with a single plot symbol.

Figure 4 shows the individual results for all biomarkers that respond to rifampicin, confirming consistent response in all individuals except for the aforementioned k(bh) value. The figure also shows the strong systematic response of the semi-quantitative measures.

**Figure 4.**
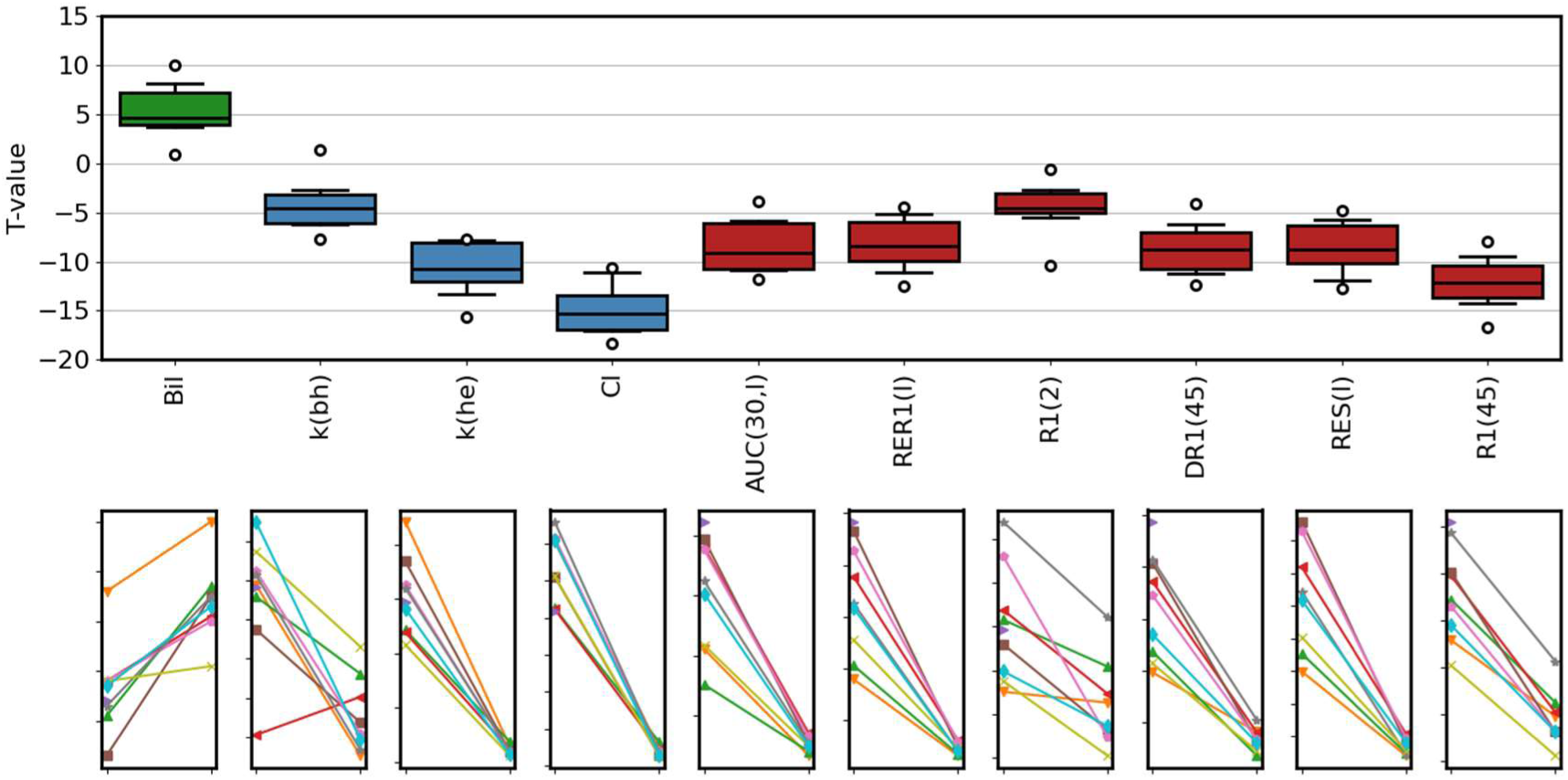
Participant-level rifampicin effect for all parameters that show a significant change in the mean value (p<0.01). The top row shows the difference between rifampicin and baseline relative to the standard error of the difference (T-value), with LFT markers in green, hepatocellular (HC) markers in blue, and semi-quantitative (SQ) markers in red. The bottom row shows the individual changes for the corresponding biomarkers. Explicit names for the biomarkers can found in tables 3, 4.

Correlation analysis at baseline (Fig 5) shows 3 distinct clusters in the MRI liver biomarkers. The hepatocellular markers k(he), k(bh) and CL are a distinct group but are not correlated; the semi-quantitative markers and the extracellular markers are all strongly correlated within the cluster. Some of the LFT’s correlate with extracellular volume but only conjugated bilirubin is correlated with k(he).

**Figure 5.**
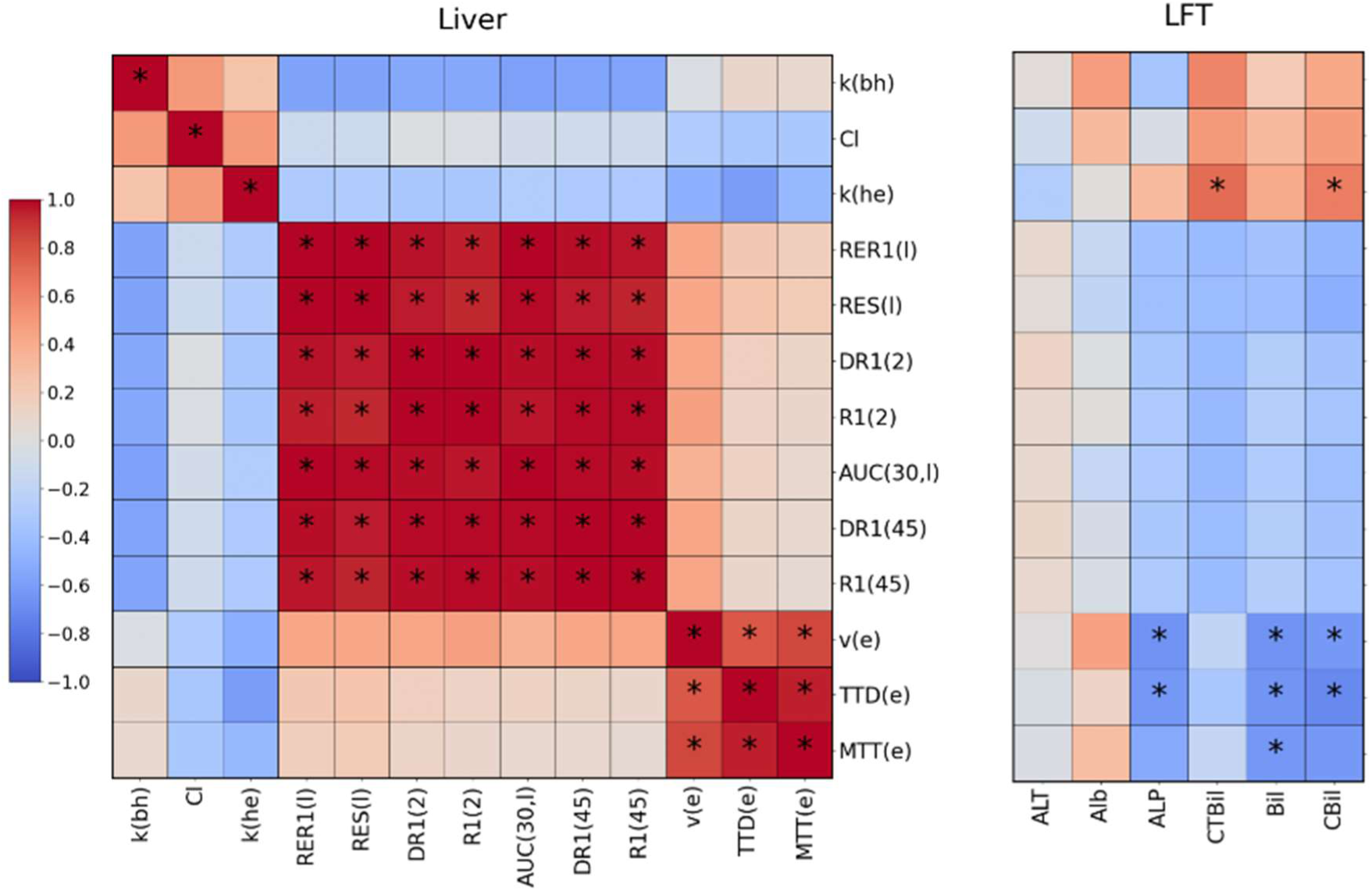
Cluster plot of correlations between liver biomarkers at baseline. The vertical axis for each panel shows the MRI liver biomarkers. The horizontal axis shows the MRI liver biomarkers (left panel) and LFT’s (right panel). Negative correlations are shown in blue and positive correlations in red, and significant correlations (p<0.01) are marked with (*). Explicit names for the biomarkers can found in tables 3, 4.

The same clusters remain visible when comparing the changes in the biomarkers after rifampicin (Fig 6). Changes in uptake and excretion rates are not significantly correlated. LFT changes show some correlations with semi-quantitative markers but not with hepatocellular markers.

**Figure 6.**
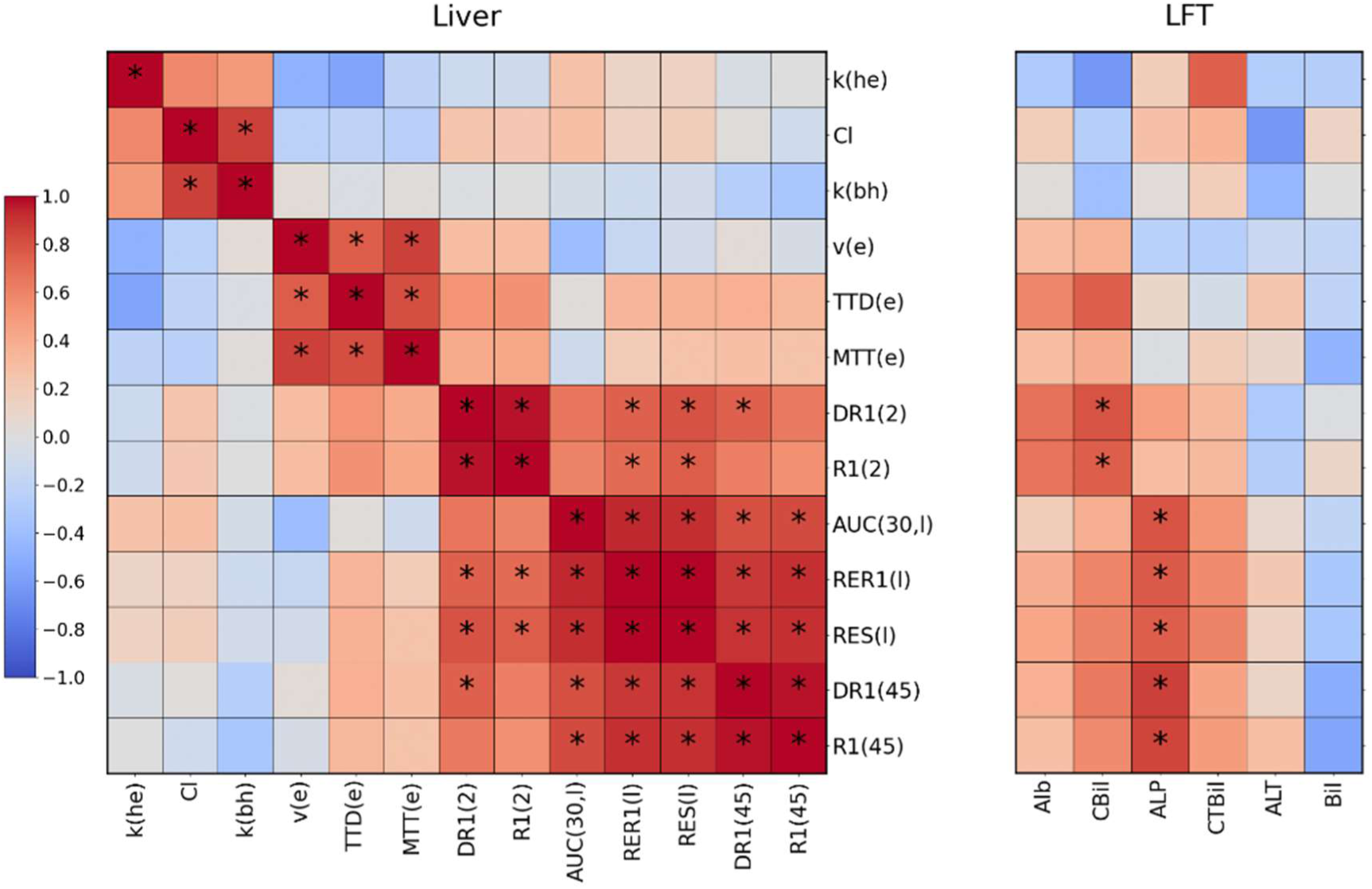
Cluster plot of correlation coefficients between changes (rifampicin – baseline) in liver biomarkers. The vertical axis in each panel shows the change in MRI liver biomarkers. The horizontal axis shows the change in MRI liver biomarkers (left panel) and change in LFT’s (right panel). Negative correlations are shown in blue and positive correlations in red, and significant correlations (p<0.01) are marked with (*). Explicit names for the biomarkers can found in tables 3, 4.

## Discussion

Effective management of DILI and liver-mediated DDI risk requires non-invasive methods to assess the effects of drugs on hepatocellular transport function. This is particularly important if modulation of transporters causes different drug exposure in the blood/plasma and liver. The aim of this study was to investigate if liver uptake and excretion rates of gadoxetate, as measured with MRI in healthy volunteers, can detect changes caused by rifampicin, a known inhibitor of OATP1B1/MRP2 transporters. Results showed a strong and consistent reduction in liver uptake and biliary excretion of gadoxetate after rifampicin administration, indicating a potential role as imaging biomarkers in drug safety assessment.

The result is consistent with findings in healthy rats [13] where rifampicin caused a 90% reduction in liver uptake and 43% in biliary excretion, in close agreement with our observations in healthy volunteers. This preclinical study also found a similar effect of cyclosporin (OATP1B1/MRP2 inhibitor) on gadoxetate, whereas other test drugs (pioglitazone, asunaprevir) showed marginal effect on excretion. The current proof-of-concept using rifampicin creates a rationale for similar validation in human participants.

Our results provide insights in the use of MRI for evaluation of transporter inhibition where changes in liver exposure differ relative to blood. Inhibition of intracellular uptake via OATP1B1 by rifampicin is stronger than the inhibition of biliary clearance via MRP2, a distinction that cannot be made with blood-based assays. Inhibition of biliary clearance is also more variable than uptake – indicating the need for a personalised approach to DDI and DILI prediction. Finally, inhibition of uptake and excretion are uncorrelated, confirming the importance of measuring both biomarkers.

The effect of rifampicin can also be detected using bilirubin data in routine LFTs. Considering the complex hepatic disposition of conjugated and unconjugated bilirubin, the effect is substantially more variable than with the MRI assay and uncorrelated to the effect on uptake and excretion – reducing the utility of LFTs for DDI assessment.

The gadoxetate-MRI assay specifically probes potential inhibitors of OATP1B1 and MRP2, allowing it to positively confirm inhibition of these transporters. However, it cannot directly detect inhibition of other hepatic transporters, such as the bile salt export pump (BSEP). Inhibition of BSEP is important because genetically inherited defects in BSEP expression can cause severe cholestatic liver injury, and because many drugs that cause human idiosyncratic DILI inhibit BSEP activity [3,25]. While gadoxetate is not a substrate of BSEP, it is possible that BSEP inhibition can be detected indirectly, for instance if cholestasis affects gadoxetate excretion.

Rifampicin effects can also be detected using semi-quantitative markers, such as post-contrast R1. These do not require modelling and are available in-line on many scanners. Similar approaches have been studied in chronic liver disease [24]. However, they do not distinguish between uptake and excretion, they cannot be compared to non-imaging assays, and their values depend on variables such as contrast agent dose. This limits their utility for DDI assessment, though they may have a role in other diagnostic questions [10].

The assay used in this study was designed for drug development and basic research, to determine whether a novel drug poses a DILI or DDI risk, or for dose setting in clinical trials. Complex experiments such as the two-scan protocol deployed in this study are realistic in this setting. In clinical applications, such as prediction of outcomes after hepatoectomy [10], a separate knowledge of billiary excretion is often less critical. In that case, a shorter protocol focused on uptake measurement may be sufficient [11].

Study limitations: The study design did not standardize preparation of the participants by controlled diet or fluid intake before the visit, and participants were not given any dietary instructions between the two scans. These factors may have contributed to the variability in rifampicin absorption and observed effects. It appears prudent for future studies to include some level of standardization in the diet and the timing of the scan.

In conclusion, gadoxetate uptake and excretion rates can quantify drug-mediated inhibition of OATP1B1/MRP2 transporters in humans, and predict the effect on liver exposure. This pavs the way for further validation using test drugs with different levels of potency, and for clinical research on drug-mediated inhibition of liver transporters in patients with impaired liver function.

## Data Availability

Signal data and numerical results are available online (see zenodo links and references in manuscript)

https://zenodo.org/records/15644122

https://zenodo.org/records/15644248

https://zenodo.org/records/15611776

https://zenodo.org/records/15616745

